# A 12-year polysomnographic study in Huntington’s: sleep problems predict disease onset and severity

**DOI:** 10.1101/2024.07.31.24311309

**Authors:** Zanna J. Voysey, Anna OG. Goodman, Lorraine Rogers, Jonathan A. Holbrook, Alpar S. Lazar, Roger A. Barker

## Abstract

Increasing evidence suggests that the sleep pathology associated with neurodegenerative diseases can in turn exacerbate both the cognitive deficits and underlying pathobiology of these conditions. Treating sleep may therefore bear significant, even disease-modifying, potential for these conditions, but how best and when to do so remains undetermined.

Huntington’s Disease (HD), by virtue of being an autosomal-dominant neurodegenerative disease presenting in mid-life, presents a key ‘model’ condition through which to advance this field. To date, however, there has been no clinical longitudinal study of sleep abnormalities in HD, and no robust interrogation of their association with disease onset, cognitive deficits and markers of disease activity. Here we present the first such study.

HD gene carriers (*n*=28) and age- and sex-matched controls (*n*=21) were studied at baseline and 10- and 12-year follow up. All HD gene carriers were premanifest at baseline, and were stratified at follow up into prodromal/manifest and premanifest groups. Sleep abnormalities were assessed through two-night inpatient polysomnography (PSG) and two-week domiciliary actigraphy, and their association was explored against i)validated cognitive and affective outcomes (Montreal Cognitive Assessment, Trail A/B task, Symbol Digit Modalities Task [SDMT], Hopkins Verbal Learning Task [HVLT], Montgomery-Asberg Depression Rating Scale [MADRS]) and ii)serum neurofilament-light (NfL) levels. Statistical analysis incorporated cross-sectional ANCOVA, longitudinal repeated measures linear models and regressions adjusted for multiple confounders including disease stage.

15 HD gene carriers phenoconverted to prodromal/early manifest HD by study completion. At follow-up, these gene carriers showed more frequent sleep stage changes (*p*=<0.001,ƞ_p_^2^=0.62) and higher levels of sleep maintenance insomnia (*p*=0.002,ƞ_p_^2^=0.52). The latter finding was corroborated by nocturnal motor activity patterns on follow-up actigraphy (*p*=0.004,ƞ_p_^2^=0.32).

Greater sleep maintenance insomnia was associated with greater cognitive deficits (Trail A *p*=<0.001,R²=0.78;SDMT *p*=0.008,R²=0.63;Trail B *p*=0.013,R²=0.60) and higher levels of NfL (p=0.015,R²=0.39).

Longitudinal modelling suggested that sleep stage instability accrues from the early premanifest phase, whereas sleep maintenance insomnia emerges closer to phenoconversion. Baseline sleep stage instability was able to discriminate those who phenoconverted within the study period from those who remained premanifest (area under curve=0.81,*p*=0.024).

These results demonstrate that the key sleep abnormalities of premanifest/early HD are sleep stage instability and sleep maintenance insomnia, and suggest that the former bears value in predicting disease onset, while the latter is associated with greater disease activity and cognitive deficits. Intervention studies to interrogate causation within this association could not only benefit patients with HD, but also help provide fundamental proof-of-concept findings for the wider sleep-neurodegeneration field.

## Introduction

Sleep abnormalities are highly prevalent across the spectrum of neurodegenerative disease^1,2^, and growing evidence suggests a deleterious, feedforward cycle between the two, in which the sleep disruption caused by neurodegeneration in turn exacerbates both the cognitive/affective features and pathophysiology of these conditions^3–5^. Sleep disturbance is known, for example, to impair executive function^6,7^, attention^8^, processing speed^9,10^ and emotional regulation^11^, and also to promote neuroinflammation^12^ and aberrant protein homeostasis^13,14^. Further, sleep is purported to play a critical role in synaptic modulation supporting memory consolidation^15^, and in glymphatic clearance of neurotoxic species such as beta-amyloid and tau from the brain^16,17^.

Treating sleep disturbance therefore bears significant, even disease-modifying, potential for neurodegenerative conditions, and the recent emergence of new sleep therapies such as orexin antagonists^18^ makes this prospect all the more feasible.

Huntington’s Disease (HD) is a fully penetrant autosomal dominant neurodegenerative disease caused by a CAG repeat expansion mutation in the *huntingtin* gene. It is characterised by a combination of motor, cognitive and psychiatric features, with disease onset (“manifest” disease) defined by the development of unequivocal motor signs, typically occurring between the ages of 35-50.

By virtue of these characteristics, HD enables sleep abnormalities to be studied longitudinally from prior to disease onset, facilitating fundamental insights into the ‘chicken’ and ‘egg’ of this feedforward cycle. Further, it allows their relation to cognitive/affective features to be studied free from the confounding effects of advanced age or comorbid health conditions. This poses a key advantage over more common neurodegenerative conditions, which occur in late-age and for which a presymptomatic or early phase can only be identified in retrospect. These same characteristics also make HD an ideal condition in which to conduct trials of new sleep therapies. Thus, HD poses a key ‘model’ condition through which to advance the sleep-neurodegeneration field.

Cross-sectional polysomnographic and actigraphic studies in manifest HD patients^19–25^ have suggested its main sleep abnormalities to comprise i)low sleep efficiency due to high levels of wake after sleep onset (ie. sleep maintenance insomnia), ii)sleep stage instability, and iii)increased light sleep with reduced slow wave sleep and rapid eye movement sleep (REM). These features appear to become more prominent with disease progression^21,24,25^. This profile mirrors that seen in Alzheimer’s and Parkinson’s Disease^26–29^, supporting the potential validity of HD as a ‘model’ condition.

Nonetheless, many of these existing studies are limited in several ways, for example a lack of habituation to polysomnography, a lack of definitive genetic diagnosis in early studies, heterogeneity of disease stage, and failure to control for medication use or affective state. Furthermore, there has been only one study to date of sleep in premanifest HD gene carriers – a study by our group^19^. This study suggested sleep stage instability and sleep maintenance insomnia to also occur at this disease stage, but without proportionate gain/loss of sleep stages.

To date, there has been no longitudinal study of sleep in HD patients, and no robust interrogation of associations between sleep abnormalities and disease onset or clinical features. The handful of studies that have considered the latter are heavily limited either by the use of subjective measures of sleep^30–32^, which are known to be unreliable in HD^20^, or by failure to control for age^33,34^.

HD also has the advantage that there is an easily-obtainable putative biomarker of disease activity in its prodromal/early stages, namely serum neurofilament light (NfL). A number of studies^35–38^ have recently demonstrated that NfL exhibits a sigmoidal trajectory in HD, with rapid increases during the late premanifest/transitional phase. Yet to date there has also been no study exploring the relationship between objective sleep abnormalities and NfL in HD.

Here, we aimed to address these knowledge gaps. We studied a cohort of HD gene carriers and age- and sex-matched healthy controls at three timepoints over a 12 year period. All HD gene carriers were premanifest at baseline; approximately half had converted to prodromal/early manifest HD by study completion. Sleep abnormalities were assessed by both inpatient polysomnography and domiciliary actigraphy, and their relationship was explored versus both cognitive/affective features and NfL levels.

Results are intended to inform the design of targeted sleep intervention trials in HD. Such studies bear potential not only to bring benefit to HD patients, but also to answer fundamental proof-of-concept questions regarding the contribution of sleep abnormalities to the presentation and progression of neurodegeneration.

## Materials and methods

### Study design

Study structure, including timing of the study subcomponents, is depicted in Figure 1. This structure reflected the influence of Covid-19 restrictions, which precluded face-to-face assessments at the time of 10-year follow up. At each timepoint, all subcomponents were undertaken within a maximum of 12 months of one another.

**Figure 1.**
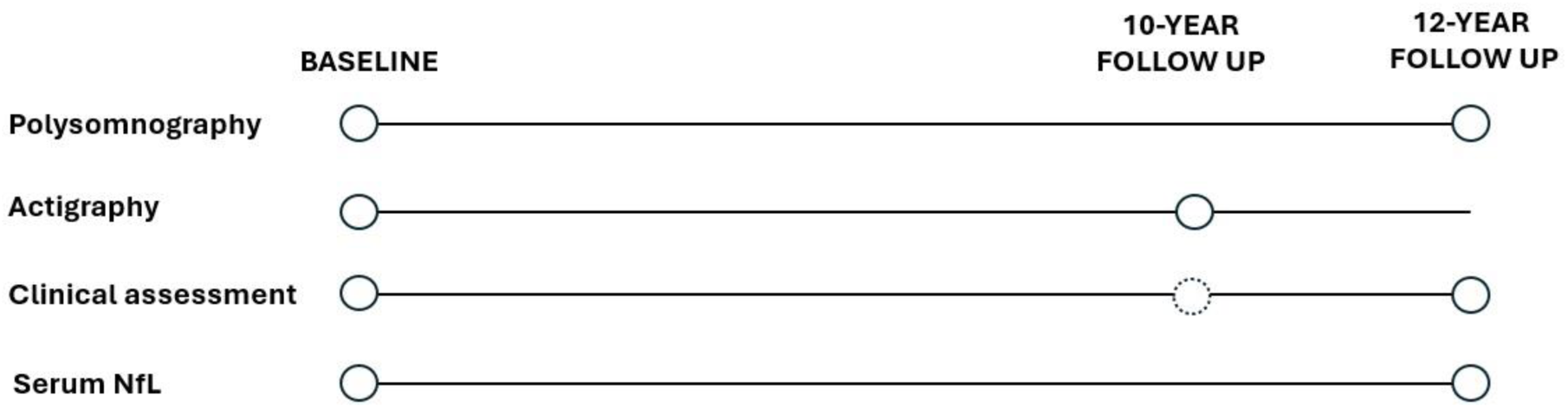
Study design. 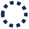 denotes where assessments were undertaken by video call in place of in-person assessment, and where Unified Huntington’s Disease Rating Scale Total Motor Score (UHDRS TMS) examination was precluded, due to Covid-19 restrictions preventing face-to-face assessments. NfL=neurofilament-light.

### Recruitment

28 patients and 22 controls were recruited in 2009-10. Approximately 50% of HD gene carriers were recruited from the Cambridge HD Clinic; the remainder self-referred from other HD clinics across the UK. Approximately 50% of controls constituted healthy partners of recruited gene carriers; the remainder were recruited by local advertisement. Ethical approval was granted by local ethics committees, and all participants provided written informed consent in accordance with the Declaration of Helsinki (REC 03/187 and 15/EE/0445).

Baseline inclusion criteria comprised i)a positive genetic test for HD, conferred by CAG repeat length ≥38 (in HD gene carriers), and ii)a Unified Huntington’s Disease Rating Scale Diagnostic Confidence Level (UHDRS DCL) of 0-1 (in HD gene carriers). The latter equates to <50% clinician confidence of signs of HD, ensuring that all HD gene carriers were premanifest at baseline. Baseline exclusion criteria comprised i)diagnosis of a sleep disorder (in controls), ii)diagnosis of any other neurodegenerative/neuroinflammatory condition or traumatic brain injury, and iii)diagnosis of a psychiatric condition bar mild-moderate anxiety or depression.

Participants were subsequently also excluded from analysis if there was i)evidence of untreated moderate sleep apnoea during polysomnography, defined as apnoea-hypopnoea index [AHI]>15^39^ (n=2; Fig.2), ii)diagnosis of any other neurodegenerative/neuroinflammatory condition or traumatic brain injury during the follow-up period (n=0), iii)diagnosis of a psychiatric condition bar mild-moderate anxiety or depression during the follow up period (n=0), or iv)night shift work or travel >2 time zones from UK <2 weeks prior to study assessments (n=0).

**Figure 2.**
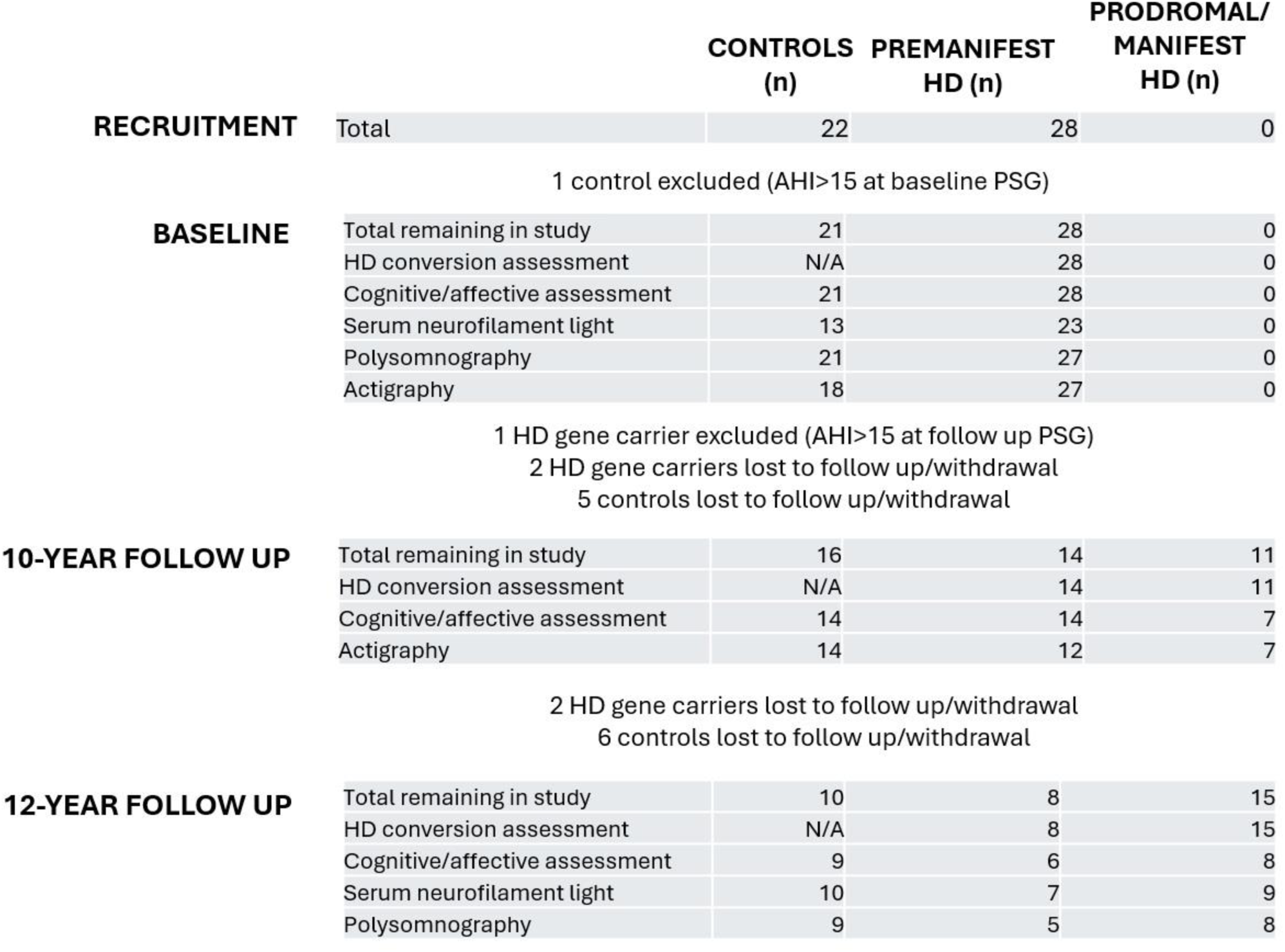
Participation and conversion flow chart: totals across study timepoints. AHI=apnoea-hypopnea index.

### Clinical assessment

The following standardised, validated clinical assessments were selected for their established sensitivity both to features of prodromal and early HD^40^ and to the effects of sleep disturbance^6,9–11^.

Cognitive/affective assessment comprised:

i) Montreal Cognitive Assessment (MoCA): test of global cognition, score 0-30, higher score equates to better performance.
ii) Trail Making Test part A (Trail A): test of attention and psychomotor speed, timed 0-180 seconds, shorter time equates to better performance.
iii) Trail Making Test part B (Trail B): test of executive function, timed 0-180 seconds, shorter time equates to better performance.
iv) Hopkins Verbal Learning Test-Revised, delayed recall component (HVLT delayed): test of learning and memory, score 0-12, higher score equates to better performance.
v) Symbol Digit Modalities Task (SDMT): test of attention and psychomotor speed, score 0-110, higher score equates to better performance
vi) Montgomery-Asberg Depression Rating Scale (MADRS): clinician rated measure of depression, range 0-60, score 7-19=mild depression, 20-34=moderate depression.

At follow up, HD gene carriers were assessed for evidence of conversion to prodromal or manifest HD according to their UHDRS total motor score (UHDRS TMS, score 0-124) and UHDRS DCL (range 0-4, score≥2 indicative of >50% confidence of signs of manifest HD). At 10-year follow up, HD gene carriers were defined as prodromal/manifest where DCL≥2^41^ (UHDRS TMS unobtainable due to Covid-19 restrictions). At 12-year follow up, this classification was made where DCL≥2 and UHDRS TMS≥4^42^.

Conversion status was also submitted where it could be ascertained from routine clinical care records and/or study participation before or after study timepoints. For example, where participants did not meet prodromal/manifest criteria at 12-year follow up, it could be reliably inferred that they had been premanifest at 10-year follow up, and conversely where participants demonstrated DCL≥2 and UHDRS TMS≥4 during clinical care assessments prior to 10-year follow up, prodromal/manifest status could be reliably inferred at 10-year follow up.

UHDRS total functional capacity scoring (UHDRS TFC, range 0-13) was also undertaken at baseline and 12-year follow up to provide a measure of disease severity (11-13=early stage; 7-10=early-mid stage HD). All UHDRS components were undertaken by certified clinicians, and all clinical assessments were undertaken blinded to sleep assessment results.

In order to mitigate against the potential effects of confounders, data was also collected during clinical assessment regarding a number of demographic factors (Table 1). Predicted years to onset of manifest HD at baseline was calculated using the Langbehn formula at 60% probability^43^; a formula based on age and CAG repeat length. Precise baseline-follow up interval was determined by months between clinical assessments. History of relevant medical comorbidity was defined as those reported by participants to disrupt to sleep on average ≥2 times per week, including perimenopausal symptoms. Use of relevant medications was defined as those for which drowsiness, sleep disorder, confusion, impaired concentration or impaired memory was listed as a common side effect in the British National Formulary 2023. History of alcohol excess was defined as >14units/week for women and >21units/week for men, for ≥3 month period. History of caffeine excess was defined as >400mg/day for ≥3 month period. For participants undertaking actigraphy, ie. a domiciliary metric, the presence of a cohabitant causing regular sleep disruption (average ≥2 times/week) was also recorded.

**Table 1.** Baseline cohort demographics. N/A=non-applicable. NS=non-significant. Group differences assessed by independent Student’s T-test or Chi square/Fisher’s exact test. *Relevant medications for HD gene carriers: carbamazepine(n=1), SSRI (n=5), SNRI(n=1), benzodiazepine(n=1), beta blocker(n=1), statin(n=1). Relevant medications for controls: SSRI(n=1), ACE inhibitor(n=1), calcium channel blocker(n=1). †Relevant medical comorbidities for HD gene carriers: post nasal drip(n=1), fibromyalgia(n=1). Where % and n are incongruent, this reflects isolated cases of missing data, polypharmacy and/or multiple comorbidities.

| Demographic variable | Controls | HD | P |
| --- | --- | --- | --- |
| N | 21 | 28 | N/A |
| Age (mean, SD) | 45.4(16.5) | 44.0(11.4) | NS |
| Sex (% male) | 42.9 | 32.1 | NS |
| CAG repeat length (mean, SD) | N/A | 41.1(2.0) | N/A |
| Predicted years to onset from baseline (mean, SD) | N/A | 18.0(10.1) | N/A |
| Years of education (mean, SD) | 15.4(3.6) | 14.7(2.4) | NS |
| In employment (%) | 88.2 | 83.3 | NS |
| Relevant medications (%)* | 14.3 | 25 | NS |
| History of alcohol excess (%) | 25 | 34.8 | NS |
| History of caffeine excess (%) | 12.5 | 43.5 | NS |
| Relevant medical comorbidities (%)† | 0 | 8.3 | NS |
| History of anxiety/depression (%) | 38.9 | 46.2 | NS |
| Cohabitant causing sleep disruption (%) | 0 | 0 | NS |

### Neurofilament-light: Meso Scale Discovery Assay

Due to the known effect of advanced age^44^ and renal impairment^45^ on serum NfL levels, participants were excluded from the NfL subcomponent where baseline age was >65 (n=3) or where there was a diagnosis of renal impairment (n=0). Extracted serum from venous blood samples was stored at −80°C until processing. NfL concentrations were determined using the Meso Scale Discovery S-PLEX Neurology Panel 1 (Human) kit according to manufacturer’s instructions, with an independent interplate control repeated across plates. All samples and standards were measured in duplicate. Plates were coated on the day of analysis and analysed using the Meso Sector 2400 Imager. Values were standardised to the independent interplate control with the lowest coefficient of variation (CV) (<2%). Samples were re-rerun where the CV exceeded the manufacturer’s recommended threshold (25%). All values fell within the dynamic range of the assay (1.7-1400pg/ml).

### Polysomnography

PSG was undertaken via an inpatient study over two consecutive nights (first night habituation, second night used for data analysis) in a light, temperature and noise controlled laboratory environment. Participants were asked to follow their typical sleep/wake routine and to refrain from caffeine intake or naps during their inpatient stay. A full clinical PSG setup was used in 10-20 EEG distribution, including tibialis anterior and submental electromyography surface electrodes. Respiratory function was assessed via pulse oximetry, nasal cannulae with pressure transducer, and thoracic respiratory effort belt. PSG signals were recorded using an Embla S7000 and visualised using RemLogic software (Embla Systems, Ontario, Canada). EEG was recorded with reference electrodes at mastoid areas (A1 and A2) with a common reference electrode at Pz. EEG signals were stored at 200Hz, with a low-pass filter at 70Hz and high-pass filter at 0.3Hz.

Sleep staging was performed in 30-second epochs according to Rechtschaffen and Kales criteria (R&K) by scorers blinded to participant identity and disease group. R&K were used as American Academy of Sleep Medicine (AASM) criteria were not established at the time of the baseline timepoint; their continued use at follow up was therefore required to facilitate comparison between timepoints, as well as between our current dataset and that derived from the same cohort in previous publications^19^. AHI and periodic limb movements were scored according to standard criteria^39,46^. Scoring of follow up PSG was cross-checked by the same individuals who had scored the baseline timepoint PSG, in order to mitigate against potential inter-rater variability.

Multiple objective PSG variables were calculated, based on standard objective sleep features used in clinical research, with the addition of three variables at follow up (Table 2). The latter reflected expanded variables of interest identified in emergent literature during the study period^19,22^. Non-proportional PSG variables occurring within sleep (arousals, sleep stage changes and limb movements) were normalised to total sleep time; those relating to wakefulness during the sleep period (awakenings and wake after sleep onset) were normalised to total time in bed. PSG data was excluded where total sleep time was <3 hours (n=1).

**Table 2.** Polysomnography outcomes. N/A=non-applicable. NS=non-significant. FU=follow up. TIB=time in bed. TST=total sleep time. Q=P value with False Discovery Rate correction applied. Group differences assessed by ANCOVA adjusted for age, sex, CAG repeat length, MADRS depression score, relevant medication use and individual interval between baseline and follow up. Values represent totals, % or mean(SD) as applicable. *p<0.05, **p<0.01, ***p<0.001 in post hoc Tukey test versus prodromal/manifest HD group. †variable added to analysis protocol at 12-year follow up. Effect size reported where p<0.05 in both pairwise post hoc assessments vs prodromal/manifest HD.

| Polysomnography variables | Baseline | | | 12-year FU | | | | $\eta_p^2$ |
| --- | --- | --- | --- | --- | --- | --- | --- | --- |
|  | Controls | HD | Q | Controls | Premanifest HD | Prodromal/Manifest HD | Q |  |
| N | 21 | 27 | N/A | 9 | 5 | 8 | N/A | . |
| TIB (hours) | 8.1(1.0) | 8.6(0.8) | NS | 8.8(0.8) | 8.4(1.6) | 9.6(1.0) | NS | . |
| TST (hours) | 6.9(1.0) | 7.2(1.2) | NS | 8.1(0.8) | 8.0(1.3) | 7.8(1.3) | NS | . |
| Sleep efficiency (%) | 85.3(7.8) | 84.1(9.8) | NS | 92.0(3.2)*** | 94.9(3.5)** | 80.6(10.8) | **0.001** | **0.56** |
| Sleep onset latency (mins) | 15.5(17.8) | 17.4(15.3) | NS | 8.0(2.9) | 5.4(3.0) | 12.5(16.2) | NS | . |
| Stage 1 (%) | 12.0(5.4) | 10.7(4.6) | NS | 8.9(3.5) | 11.3(5.3) | 12.6(5.2) | NS | . |
| Stage 2 (%) | 51.7(5.9) | 51.7(8.5) | NS | 43.3(3.5) | 41.5(6.9) | 37.8(7.1) | NS | . |
| Slow wave sleep (%) | 17.0(7.2) | 17.4(8.1) | NS | 23.1(5.2) | 19.1(4.4) | 28.3(6.6) | NS | . |
| REM sleep (%) | 19.3(6.0) | 21.0(5.0) | NS | 24.7(5.8) | 31.3(3.3)** | 20.9(5.2) | **0.03** | . |
| Wake after sleep onset (mins/hour) | 6.8(3.8) | 6.4(3.8) | NS | 3.9(2.0)** | 2.4(2.0)*** | 10.3(5.6) | **0.005** | **0.52** |
| Awakenings/hour | 1.2(0.7) | 1.3(0.7) | NS | 1.6(0.7) | 1.2(0.4) | 2.2(1.1) | NS | . |
| Limb movement arousals/hour | 2.8(1.1) | 3.4(1.4) | NS | 5.7(3.3) | 5.7(1.7) | 5.9(2.9) | NS | . |
| Sleep stage changes/hour | 25.6(6.7) | 26.2(8.0) | NS | 25.1(6.9)** | 23.6(3.8)*** | 34.0(6.1) | **0.001** | **0.62** |
| Limb movements/hour† | . | . | . | 9.7(5.2) | 11.2(2.0) | 9.0(4.5) | NS | . |
| Periodic limb movements/hour† | . | . | . | 3.2(3.8) | 1.7(2.2) | 1.5(1.9) | NS | . |
| Arousals/hour† | . | . | . | 13.0(3.9) | 12.6(4.8) | 18.6(4.3) | NS | . |

### Actigraphy

Actigraphy was undertaken via 14 consecutive day/night domiciliary recordings during which participants wore an actiwatch (MotionWatch8, CamNTech, Cambridge, UK) continuously on their non-dominant wrist. Participants were instructed to follow their habitual sleep/wake patterns, and to complete a daily sleep diary to document recording anomalies (for example, non-representative rest-activity patterns due to transient intercurrent illness, or where there was delay in re-siting an actiwatch following bathing): such data periods were excluded (Table 3).

**Table 3.**
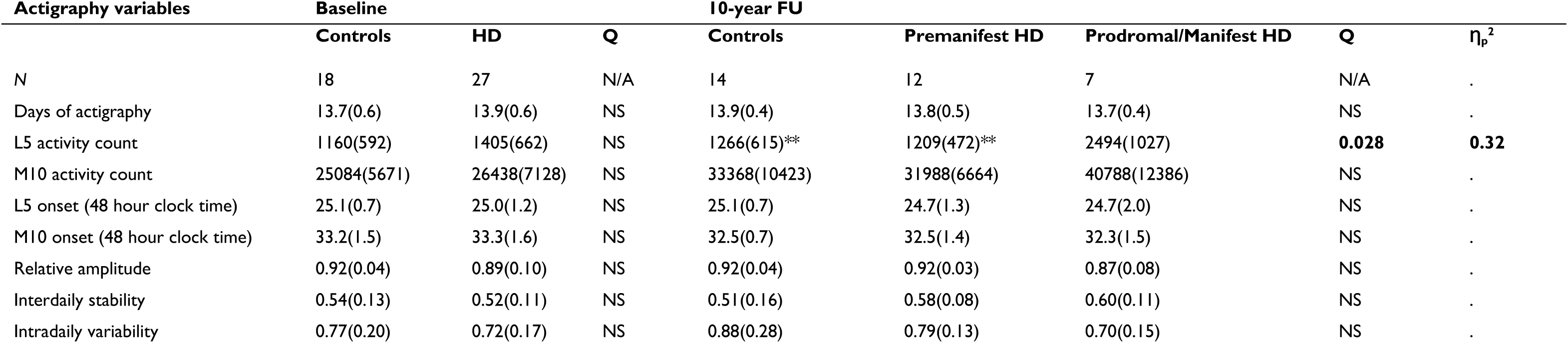
Actigraphy outcomes. N/A=non-applicable. NS=non-significant. FU=follow up. Q=p value following False Discovery Rate correction. Group differences assessed by univariate general linear models adjusted for age, sex, CAG repeat length, MADRS depression score, relevant medication use and individual interval between baseline and follow up. Values represent totals or mean(SD) as applicable. *p<0.05, **p<0.01, ***p<0.001 in post hoc Tukey test versus prodromal/manifest HD group. Effect size reported where p<0.05 in both pairwise post hoc assessments vs prodromal/manifest HD.

| Actigraphy variables | Baseline | | | 10-year FU | | | | $\eta_p^2$ |
| --- | --- | --- | --- | --- | --- | --- | --- | --- |
|  | Controls | HD | Q | Controls | Premanifest HD | Prodromal/Manifest HD | Q |  |
| N | 18 | 27 | N/A | 14 | 12 | 7 | N/A | . |
| Days of actigraphy | 13.7(0.6) | 13.9(0.6) | NS | 13.9(0.4) | 13.8(0.5) | 13.7(0.4) | NS | . |
| L5 activity count | 1160(592) | 1405(662) | NS | 1266(615)** | 1209(472)** | 2494(1027) | **0.028** | **0.32** |
| M10 activity count | 25084(5671) | 26438(7128) | NS | 33368(10423) | 31988(6664) | 40788(12386) | NS | . |
| L5 onset (48 hour clock time) | 25.1(0.7) | 25.0(1.2) | NS | 25.1(0.7) | 24.7(1.3) | 24.7(2.0) | NS | . |
| M10 onset (48 hour clock time) | 33.2(1.5) | 33.3(1.6) | NS | 32.5(0.7) | 32.5(1.4) | 32.3(1.5) | NS | . |
| Relative amplitude | 0.92(0.04) | 0.89(0.10) | NS | 0.92(0.04) | 0.92(0.03) | 0.87(0.08) | NS | . |
| Interdaily stability | 0.54(0.13) | 0.52(0.11) | NS | 0.51(0.16) | 0.58(0.08) | 0.60(0.11) | NS | . |
| Intradaily variability | 0.77(0.20) | 0.72(0.17) | NS | 0.88(0.28) | 0.79(0.13) | 0.70(0.15) | NS | . |

Actiwatches comprised triaxial accelerometers recording peak intensity of movement each second, sampled at 50Hz, expressed as an activity count in 30 second epochs. Actograms were analysed using MotionWare software (CamNTech, Cambridge, UK) according to previously published non parametric circadian rhythm analysis algorithms^20^. In brief, the time of onset and mean activity levels during the lowest 5 hours of activity (ie. reflecting nocturnal sleep, L5), and highest 10 hours of activity (M10), are calculated for each 24 hour period and averaged over the recording period. L5 and M10 are then combined to generate a measure of the relative amplitude of rest-activity levels (RA). Interdaily stability is calculated to provide an indication of regularity of rest-activity patterns across days (scale 0-1, higher indicating greater stability), while intradaily variability is calculated to provide an indication of consolidation of rest versus activity within days (scale 0-2, values>1 indicating abnormal fragmentation). By adopting this approach, actigraphy data did not require adjustment according to participant-estimated bed/wake times. Since such estimates are liable to inaccuracy/interindividual variability in precision, this maximised the reliability of results.

### Statistical analysis

Statistical analysis was undertaken in SPSS v29.0.0 (IBM, Armonk, NY, USA). The threshold for statistical significance for all analyses was p<0.05 (two-tailed).

Outlier datapoints were defined as those falling >3SDs from group means and causing skew from normal distribution and were excluded. This applied to <0.5% (n=8) datapoints across the entire dataset.

Cross-sectional group differences were assessed by one-way analysis of covariance (ANCOVA). Longitudinal group differences were assessed by repeated measures general linear models and linear mixed models. Associations between sleep variables and clinical outcomes/NfL levels were assessed by multivariate linear regression.

Variables included within and/or residuals resulting from parametric models were assessed for normal distribution by Shapiro-Wilk test and QQ plot, with variables transformed where necessary.

Covariate adjustment was based on both the a priori strength of effect of known biological confounders (eg. age, sex, depression and relevant medication use) and the presence/absence of group differences in confounders within our dataset. Given i)the high number of potential confounders relevant to sleep and cognitive data, ii)the necessarily low number of observations given the rarity of the condition and extended time period of follow up, and iii)the likelihood of combinatorial effects and collinearity between relevant covariates, we judged combined backward and forward selection of covariates, alongside assessment for collinearity by variance inflation factor, to represent the most parsimonious and stringent approach to our dataset^47^. Specifically, within ANCOVAs and longitudinal linear models, we adjusted for age, sex, CAG repeat length, MADRS depression score, relevant medication use, and baseline-follow up interval time via stepwise backward elimination, with age and sex then resubmitted (where previously eliminated) by forward selection. This approach was then modified for regression analyses between sleep metrics and clinical measures among HD gene carriers, since the baseline-follow up interval was no longer relevant, but adjustment for disease stage became important. In these analyses, we therefore replaced baseline-follow up interval with predicted years to disease onset at baseline. Since the latter is derived from a formula based on CAG repeat length^43^, CAG repeat length was omitted from the stepwise protocol to avoid collinearity.

Group differences in polysomnography and actigraphy metrics were adjusted for multiple comparisons by Benjamini-Hochberg correction^48^ (False Discovery Rate <0.05), given the high number of derived variables.

Effect sizes for group differences were expressed as partial eta square (ƞ_p_^2^) or Cohen’s f^2^, as applicable.

Receiver operating characteristic (ROC) curve and area under the curve (AUC) analyses were used to explore the ability of sleep variables to discriminate phenoconversion patterns among HD gene carriers over the study period.

### Data availability

The data that support the findings of this study are available on request from the corresponding author. The data are not publicly available due to their containing information that could compromise the privacy of research participants.

## Results

### Demographics

The baseline cohort comprised 21 controls (43% male, age 45.4±16.5) and 28 premanifest HD gene carriers (32% male, age 44.0±11.4). HD gene carriers were on average 18.0±10.1 years from predicted conversion to manifest HD at baseline (CAG range 38-46). No participant had a diagnosed sleep disorder at baseline. The distribution between controls, premanifest HD and prodromal/manifest HD gene carriers was 21:28:0 at baseline, 16:14:11 at 10-year follow up, and 10:8:15 at 12-year follow up. Figure 2 details precise rates of participation in each study subcomponent, and cases of exclusion and loss to follow up/withdrawal.

HD gene carriers and controls did not differ with respect to confounding factors at baseline (Table 1). Similarly, groups did not differ in any of these factors at 10-year and 12-year follow up, other than with respect to precise baseline-follow up interval, and, as would be expected, higher CAG repeats and lower predicted years to onset from baseline among HD gene carriers who converted to prodromal/manifest HD (Supplementary Tables 1-4). These factors were therefore included in covariate adjustment (see Methods). There was also no evidence of significant withdrawal bias in gene carriers across the study period (Supplementary Tables 5-6).

### Clinical assessment

At baseline, HD gene carriers exhibited no differences from controls with respect to cognitive/affective measures (Supplementary Table 7). However, by 10-year follow up, HD gene carriers who had converted to prodromal/manifest disease exhibited impaired attention and psychomotor speed (Trail A *F*(2,30)=4.78, *p*=0.016, ƞ_p_^2^=0.24; SDMT *F*(2,30)=16.39, *p*=<0.001, ƞ_p_^2^=0.52), executive function (Trail B *F*(2,31)=16.64, *p*=0.002, ƞ_p_^2^=0.52), and learning/memory (HVLT delayed *F*(2,27)=19.15, *p*=<0.001, ƞ_p_^2^=0.59) compared both to controls and HD gene carriers who had remained premanifest (Supplementary Table 7). These deficits were also evident in group differences at 12-year follow up, with the exception of learning/memory (HVLT delayed) which no longer met statistical significance, likely due to the reduced cohort size (Supplementary Table 7).

MADRS depression scores did not differ between groups at any timepoint, and only one participant met criteria for moderate depression at any follow up timepoint (HD gene carrier; score=25 at 12-year follow up).

UHDRS TFC among prodromal/manifest HD gene carriers at 12-year follow up was 10.9±2.7, indicating that these participants remained in the early stages of disease by study completion (Supplementary Table 7).

To explore the possible influence of video vs in-person clinical assessment (due to Covid-19 restrictions), we compared results at 10- and 12-year follow up in participants who had undergone both forms of assessment (*n*=20). There were no significant differences in any clinical assessment result (Supplementary Table 8). To consider the possible impact of the absence of UHDRS TMS score in conversion assessments at 10-year follow up (again due to Covid-19 restrictions), we also analysed discrepancies between 10- and 12-year follow up group classification. In no instance was a gene carrier scored as premanifest at 10-year follow up but manifest at 12-year follow up.

### Neurofilament-light

At baseline, NfL concentrations did not differ significantly between HD gene carriers and controls, but were elevated among prodromal/manifest HD gene carriers at 12-year follow up compared to both controls and gene carriers who had remained premanifest (Supplementary Table 7, *F*(2,23)=14.7, *p*=0.003, ƞ_p_^2^=0.51). NfL levels between controls and premanifest HD gene carriers at 12-year follow up were not significantly different (*F*(1,15)=0.58, *p*=0.88).

The cognitive and NfL profiles seen in our study were therefore concordant with that published data in HD patients^40,49^ other than a relatively low prevalence of depression^50^.

### Polysomnography

There were no differences in PSG variables between HD gene carriers and controls at baseline (Table 2). However, at 12-year follow up, HD gene carriers who had converted to prodromal/manifest disease exhibited a number of abnormalities compared to both controls and premanifest HD gene carriers: an increase in the frequency of sleep stage changes (SSC *F*(2,17)=13.65, p=<0.001, ƞ_p_^2^=0.62), a reduction in sleep efficiency (SE *F*(2,18)=11.47, *p*=<0.001, ƞ_p_^2^=0.56), and high levels of wake after sleep onset (WASO *F*(2,18)=9.62, *p*=0.002, ƞ_p_^2^=0.52) (Table 2, Fig. 3). Correlation analysis demonstrated that the decline in sleep efficiency in these individuals was due to elevated WASO (*p*=<0.001, ρ=-0.97) rather than sleep onset latency (SOL *p*=0.25, ρ=-0.35). Total WASO indicated that 88% of prodromal/manifest HD gene carriers met clinical thresholds for sleep maintenance insomnia (WASO>30minutes^51^), compared to only 20% of premanifest HD gene carriers.

**Figure 3.**
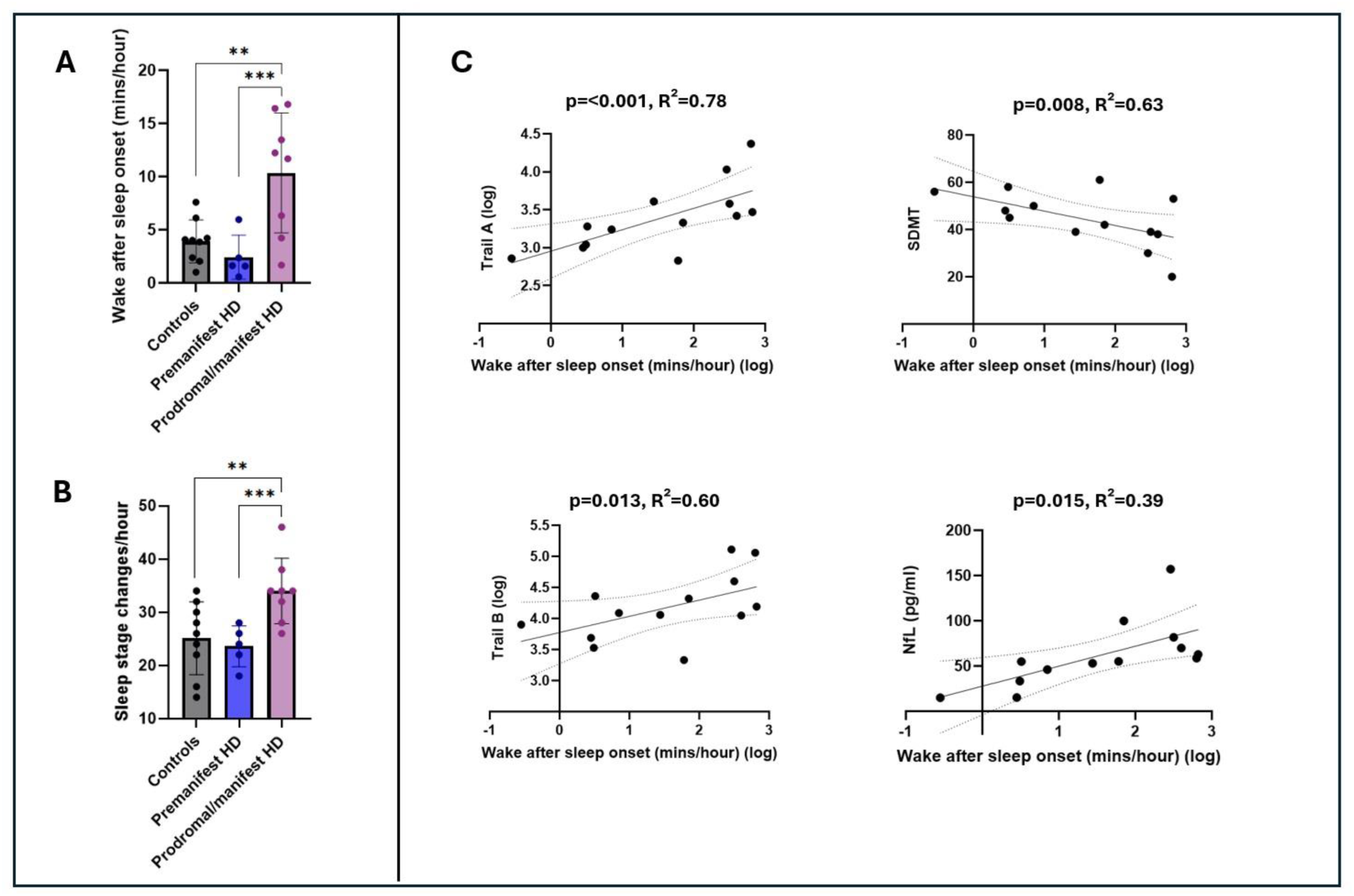
Polysomnography outcomes and associations with clinical markers. Group differences in wake after sleep onset **(A)** and sleep stage changes **(B)** on polysomnography at 12-year follow up, assessed by ANCOVA adjusted for age, sex, CAG repeat length, MADRS depression score, relevant medication use and individual interval between baseline and follow up. Error bars=±1SD. *p<0.05, **p<0.01, ***p<0.001 in post hoc Tukey test. **(C)** Association between wake after sleep onset and clinical measures in HD gene carriers at 12-year follow up, assessed by linear regression adjusted for age, sex, predicted years to onset at baseline, MADRS depression score and relevant medication use.

Prodromal/manifest HD gene carriers also exhibited reduced REM sleep compared to premanifest HD gene carriers, but this did not meet significance versus controls (Table 2).

Supplementary Figure 1 provides illustrative examples of PSG hypnograms from the three groups.

Longitudinal modelling indicated significant differences in the dynamics of these variables over the study period: HD gene carriers who converted to prodromal/manifest disease gained in WASO over the study period, whereas WASO levels were in fact lower in those who remained premanifest and controls at follow up compared to baseline (Fig. 4). Consistent with this, longitudinal modelling revealed a significant group*time interaction for WASO (*F*(2,18)=5.17, *p*=0.017, ƞ_p_^2^=0.37). By contrast, there was no such significant group*time interaction for SSC: there were similar patterns of increase in SSC among gene carriers across the study period, irrespective of whether they remained premanifest or converted to prodromal/manifest HD by study completion, compared to stable levels among controls (Fig. 4, *F*(2,15)=0.87, *p*=0.86).

**Figure 4.**
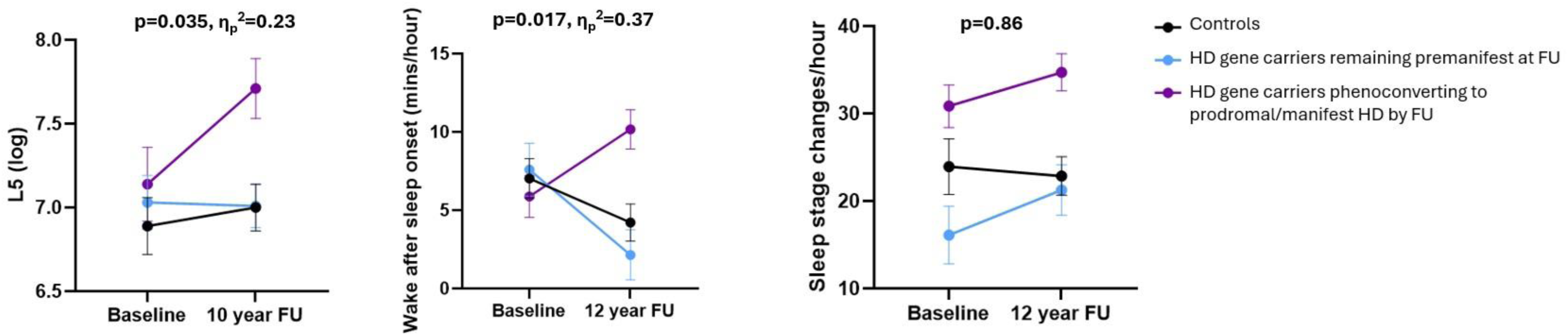
Longitudinal modelling of actigraphy L5 **(left)**, PSG wake after sleep onset **(middle)** and PSG sleep stage changes **(right)**. Values represent estimated marginal means±1SEM. P values represent group*time interaction significance in repeated measures general linear models adjusted for age, sex, CAG repeat length, MADRS depression score, relevant medication use and individual interval between baseline and follow up. FU=follow up.

To mitigate against the potential influence of loss to follow up, we also undertook longitudinal modelling of WASO using a linear mixed model adjusted for the same covariates. The group*time interaction remained statistically significant following this (*p*=0.002, Cohen f^2^=0.53).

We then assessed for associations between these sleep abnormalities at 12-year follow up and cognitive/affective features or NfL levels in HD gene carriers. We found no such association with respect to SSC. However, greater WASO was associated with higher NfL levels (*p*=0.015, R²=0.39) as well as greater deficits in attention and psychomotor speed (Trail A *p*=<0.001, R²=0.78; SDMT *p*=0.008, R²=0.63) and executive function (Trail B *p*=0.013, R²=0.60) at 12-year follow up (Fig. 3). This association survived adjustment for multiple confounders including disease stage, relevant medication use and depression scores (Fig. 3). There was no association between WASO and MADRS depression scores (*p*=0.59) or learning/memory (HVLT delayed, *p*=0.14).

### Actigraphy

At baseline, HD gene carriers did not differ from controls with respect to any actigraphy variable (Table 3). However, at 10-year follow up, gene carriers who had converted to prodromal/manifest disease exhibited elevated levels of nocturnal motor activity (L5) compared to both premanifest gene carriers and controls (Table 3, Fig. 5, *F*(2,29)=6.75, *p*=0.004, ƞ_p_^2^=0.32). This was also reflected longitudinally, with a significant group*time interaction evident for L5 (Fig. 4, *F*(2,26)=3.83, *p*=0.035, ƞ_p_^2^=0.23).

**Figure 5.**
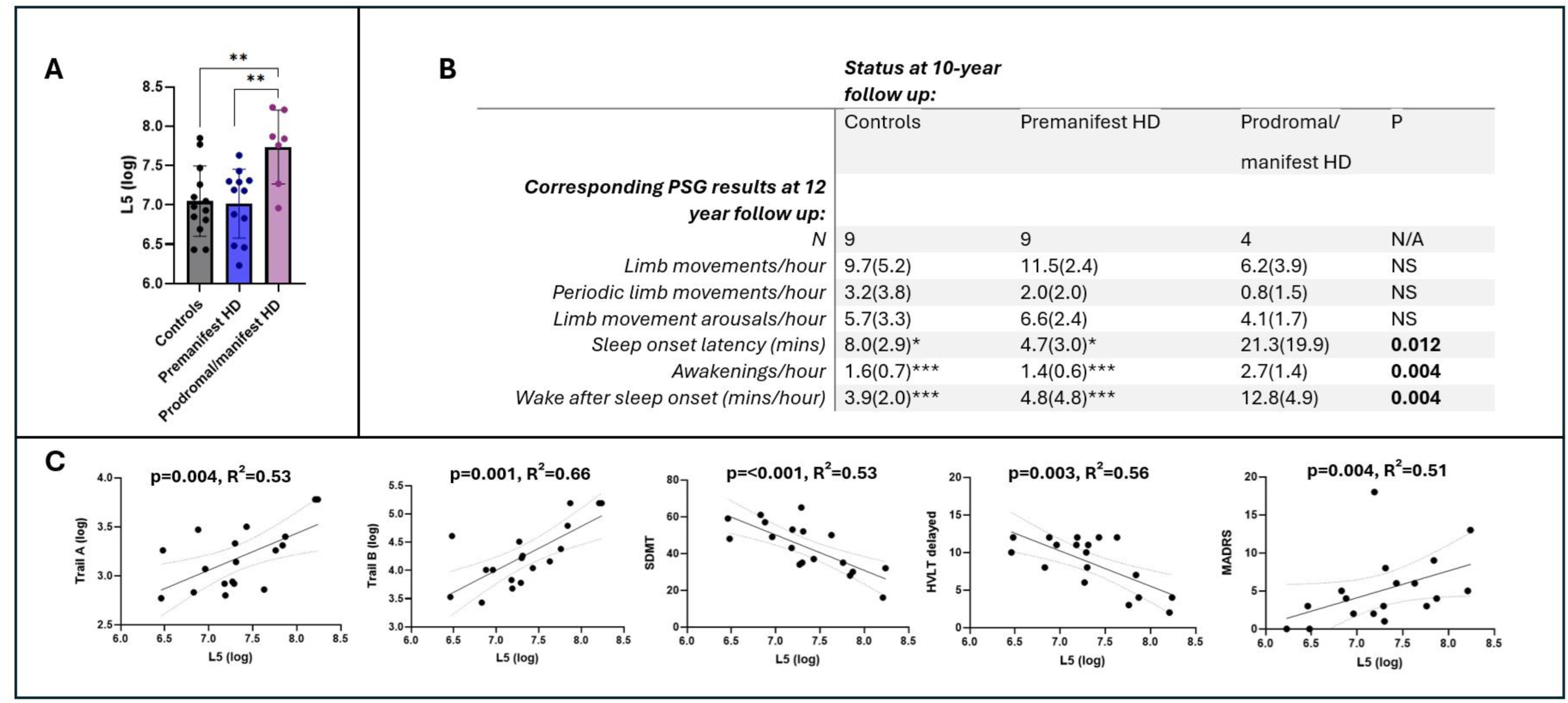
Actigraphy outcomes and associations with clinical markers. **(A)** Group differences in sleep period activity (L5) on actigraphy at 10-year follow up, assessed by ANCOVA adjusted for age, sex, CAG repeat length, depression score, relevant medication use and individual interval between baseline and follow up. Error bars=±1SD. **(B)** Conversion status at 10-year follow up in relation to relevant polysomnography variables at 12-year follow up. *p<0.05, **p<0.01, ***p<0.001 in post hoc Tukey test versus prodromal/manifest HD group. **(C)** Association between L5 and clinical measures in HD gene carriers at 10-year follow up, assessed by linear regression adjusted for age, sex, predicted years to onset at baseline, MADRS depression score and relevant medication use.

By chance, all HD gene carriers who had converted to prodromal or manifest HD at 10-year follow up were female (Supplementary Table 2). Therefore, to further assess the possible influence of sex on our results beyond covariate adjustment, we also conducted female-only analysis across groups, in which the difference in L5 between groups at 10-year follow up remained statistically significant (*F*(2,17)=4.90, *p*=0.021, ƞ_p_^2^=0.37).

Since actigraphy purely reports movement, this finding could have reflected motor activity in sleep (for example, repositioning movements, periodic limb movements, limb movement arousals or REM behaviour sleep disorder) or motor activity during nocturnal wakeful periods (for example, due to movement during brief awakenings, sleep onset insomnia or sleep maintenance insomnia). To interrogate this, we cross-referenced HD gene carriers’ actigraphy results at 10 years with their PSG results at 12 years. This revealed that individuals with high nocturnal movement on actigraphy (ie. those who were prodromal/manifest during 10-year actigraphy) exhibited higher rates of wakefulness on their subsequent PSG (SOL *F*(2,19)=5.60, *p*=0.012,ƞ_p_^2^=0.37; awakenings *F*(2,18)=7.30, *p*=0.004,ƞ_p_^2^=0.44; WASO *F*(2,19)=7.70, *p*=0.004, ƞ_p_^2^=0.45), but showed no difference with respect to movements within sleep (Fig. 5). Thus, the elevated nocturnal movement (L5) result more likely represented motor activity during periods of nocturnal wakefulness than movements in sleep. As groups did not differ with respect to daytime activity levels (M10, Table 3), it is unlikely that chorea during nocturnal wakefulness made a substantial contribution to this nocturnal motor activity.

We then assessed for associations between L5 and cognitive/affective features among HD gene carriers at 10-year follow up. Paralleling the WASO findings on PSG, higher L5 was associated with greater deficits in attention and psychomotor speed (Trail A *p*=0.004, R²=0.53; SDMT *p*=<0.001, R²=0.53) as well as executive function (Trail B *p*=0.001, R²=0.66) (Fig. 5). In addition, higher L5 was also associated with greater deficits in learning/memory (HVLT delayed *p*=0.003, R²=0.56) and higher depression scores (MADRS *p*=0.004, R²=0.51) (Fig. 5). These associations between L5 and cognitive deficits survived adjustment for disease stage, relevant medication use, depression scores and multiple other confounders (see Methods).

### Retrospective baseline analysis

We then explored whether sleep profiles varied by proximity to disease onset, or bore predictive value in estimating this. To achieve this, we retrospectively stratified the baseline cohort of HD gene carriers according to clinical outcome at study completion: those who had converted to prodromal/manifest disease by study completion were termed <12 years from conversion at baseline, whereas those who remained premanifest were termed >12 years from conversion at baseline.

When considering baseline sleep variables for these groups, HD gene carriers <12 years from conversion exhibited elevated SSC compared to those >12 years from conversion (mean 29.2±8.5 versus 20.7±5.8 changes/hour, *F*(1,17)=12.32, *p*=0.003, ƞ_p_^2^=0.42). No other sleep variable, including WASO and L5, exhibited group differences. Thus, our results suggest that sleep stage instability develops earlier than sleep maintenance insomnia in premanifest HD gene carriers.

In support of this, ROC curve analysis revealed that baseline SSC showed good ability to discriminate HD gene carriers who phenoconverted to prodromal/manifest disease during the study period from those who remained premanifest (AUC=0.81, *p*=0.024), with a SSC cut off score of 23.2 changes/hour exhibiting 69% sensitivity and 71% specificity in determining this. Baseline WASO, by contrast, did not exhibit parallel efficacy (AUC=0.51, *p*=0.942). At follow up, both SSC and WASO exhibited excellent ability to discriminate prodromal/manifest from premanifest gene carriers (SSC AUC=0.95, *p*=0.008; WASO AUC=0.93, *p*=0.013). A WASO cut off score of 3.3mins/hour and a SSC cut off score of 27.0 changes/hour each exhibited 88% sensitivity and 80% specificity in identifying this (Supplementary Fig. 2).

## Discussion

Here, we present the first clinical study to examine the longitudinal dynamics of sleep abnormalities in HD, and the first robust interrogation of the associations between these sleep abnormalities versus disease onset, clinical features and markers of disease activity.

We show that, although total sleep time and sleep stage proportions are preserved, patients with prodromal/early manifest HD exhibit less consolidated sleep, characterised by high levels of wake after sleep onset (sleep maintenance insomnia) and frequent sleep stage changes. Sleep stage instability appears to accrue gradually from early within the premanifest phase, whereas sleep maintenance insomnia appears later in this transition to manifest disease.

We then show, for the first time, that the more HD patients experience sleep maintenance insomnia, the worse their cognitive impairment (attention, psychomotor speed and executive function) and the higher their markers of disease activity (NfL). Since these associations survived adjustment for multiple confounders including disease stage, relevant medication use and depression scores, this suggests that sleep maintenance insomnia is not merely an epiphenomenon of disease progression or affective state, but potentially an independent contributor to these measures. This therefore raises the possibility that treating sleep maintenance insomnia in prodromal/early HD could improve cognitive outcomes and/or disease progression. This is significant, given that i)we currently have no proven therapies that achieve this for HD, and ii)we now have new agents, such as orexin antagonists, with which to treat sleep maintenance insomnia. Adding weight to this notion, moreover, cognitive and survival benefit has been observed in response to sleep interventions in transgenic mouse models of HD, including with orexin antagonists^52,53^.

We also show, for the first time, that sleep features may carry value in predicting proximity to manifest HD, since levels of sleep stage instability were able to discriminate gene carriers who phenoconverted during the subsequent 12 years from those who did not. Importantly, this mirrors findings in more common neurodegenerative diseases, where sleep abnormalities constitute a key component of the disease prodrome^54,55^.

Our identified sleep phenotype is consistent with our previous cross-sectional study^19^. Nonetheless, it should be acknowledged that there was overlap of participants between these two studies: independent replication studies are therefore needed. Furthermore, the fact that we did not find gene carrier versus control group differences in sleep variables at baseline is at odds with our previous paper^19^, where sleep stage instability and sleep maintenance insomnia was seen versus controls in premanifest HD patients predicted to be >18 years from disease onset. This may reflect the lower cohort size of our current study compared to our previous one. Nonetheless, our findings regarding sleep stage instability in retrospective baseline analysis is in keeping with sleep pathology being present in HD many years prior to conversion to manifest disease.

Our findings of a relationship between sleep maintenance insomnia and impaired attention, psychomotor speed and executive function is in line with evidence from healthy individuals^6–10^. This is particularly the case given that sleep continuity has been found to bear greater influence on cognitive performance than total sleep time^9,56^. To date, there have been few studies assessing these associations in the setting of neurodegeneration, but the limited evidence available would also suggest their presence in Alzheimer’s and Parkinson’s^4,57,58^. Likewise, studies in Alzheimer’s and Parkinson’s have found correlation between sleep disturbance and NfL levels^59,60^. It is perhaps surprising that we did not find stronger evidence of an association between sleep disturbance and depression: this is likely a corollary of both sample size and relatively low rates of depression in our cohort^50^. However, this profile within our cohort argues against our findings being attributable to depression.

Our study is notable for a number of strengths. The 12 year timespan of the study is, for example, not only the longest study of this type in HD, but also enabled us to perform group stratification based on actual rather than predicted conversion outcomes. This is an advantage over the majority of HD studies in premanifest gene carriers, which base stratification on predicted proximity to disease onset; a prediction known to be of limited accuracy^61^. Moreover, we controlled for a large number of potential confounding factors, many of which have been overlooked in previous studies of sleep in neurodegenerative conditions. Above all, it is striking that our PSG and actigraphy results provided independent corroboration of one another. By incorporating both of these approaches, we were able to counterbalance high-precision but short-duration sleep data recorded in an unnatural environment (PSG), versus lower-precision but long-duration data in an ecologically valid environment (actigraphy). The fact that we found parallel findings on both of these approaches therefore adds weight to our results.

Nonetheless, our study has a number of limitations. Firstly, while our cohort is sizeable given the rarity of HD as a condition and the timeframe of follow-up, absolute numbers of participants undertaking some study subcomponents, particularly at follow up PSG, are small. The lack of a study timepoint midway through our study period also limits the precision with which we can estimate the longitudinal dynamics of HD sleep abnormalities. For example, we cannot infer at what point in the 12 years prior to conversion to prodromal/manifest disease HD gene carriers typically develop sleep maintenance insomnia – i.e. whether this typically occurs in the decade prior to, or more concomitantly with, motor manifestation of HD. Future studies should consider this. Secondly, as no HD gene carriers reached advanced disease by study completion, our findings also cannot be extrapolated to later disease stages. Indeed, cross-sectional studies predict that the profile of sleep abnormalities may evolve across the natural history of HD, with loss of REM and slow wave sleep likely becoming more prominent in advanced disease^19,22,25^. Thirdly, the reduction in WASO between baseline and follow up among controls and premanifest HD gene carriers suggests that participants may not have been fully habituated to PSG at baseline. Fourthly, while we undertook multiple steps to mitigate against the effects of Covid-19 restrictions (see Results: clinical assessments), their possible impacts must also be considered, particularly with regard to conversion assessment and the absence of NfL measurement at 10-year follow up. Moreover, NfL remains an exploratory biomarker of disease activity in HD, and future studies would be enhanced by the incorporation of additional measures such as caudate thinning on volumetric MRI^62^.

Future studies would also be enriched by incorporating mechanistic metrics, to explore the biological basis for these observed sleep abnormalities. Longitudinal melatonin secretion profiles within HD gene carriers will be the subject of a forthcoming publication from our group, but additional longitudinal studies looking at, for example, orexin levels and hypothalamic/brainstem pathology in HD patients^63^ versus sleep abnormalities would also help to inform intervention studies.

Above all, this study is limited as causation clearly cannot be inferred from the observed associations. Intervention studies are therefore now warranted to probe whether treating sleep maintenance insomnia in prodromal/early HD can improve cognition and/or disease progression. Such a study would not only benefit HD patients, but is also ideally placed to provide fundamental proof-of-concept findings regarding the contribution of sleep abnormalities to the features and pathobiology of neurodegeneration.

## Acknowledgements

We would like to thank all the participants, and their families, for their contribution to this study, particularly given the longterm commitment this required. We thank Dr M. Camacho for advising on statistical analysis of the data. We acknowledge Prof. E. Wild in our use of Huntington’s disease conditional onset probability calculator based on the Langbehn formula.

