## Supplemental tables and figures for "A 12-year polysomnographic study in Huntington’s: sleep problems predict disease onset and severity"

| **10-year follow up cohort demographics** | *Controls* | *Premanifest HD* | *Prodromal/manifest HD* | *P* |
|---|---|---|---|---|
| *N* | 16 | 14 | 11 | N/A |
| *Baseline:10 year follow up interval (years) (mean, SD)* | 9.5(1.2) | 10.4(0.4) | 9.8(0.8) | **0.020** |
| *Age (mean, SD)* | 58.6(17.1) | 52.4(12.0) | 57.6(11.6) | NS |
| *Sex (% male)* | 43.8 | 42.9 | 9.1 | NS |
| *CAG repeat length (mean, SD)* | N/A | 40.4(1.7) | 41.8(2.2) | NS |
| *Predicted years to onset from baseline (mean, SD)* | N/A | 22.9(10.5) | 12.1(6.5) | **0.007** |
| *Years of education (mean, SD)* | 15.4(3.6) | 13.7(2.1) | 15.6(2.2) | NS |
| *In employment (%)* | 56.3 | 64.3 | 20.0 | NS |
| *Relevant medications (%)** | 21.4 | 30.8 | 28.6 | NS |
| *History of alcohol excess (%)* | 25 | 46.2 | 11.1 | NS |
| *History of caffeine excess (%)* | 12.5 | 46.2 | 33.3 | NS |
| *Relevant medical comorbidities (%)†* | 0.0 | 15.4 | 20.0 | NS |
| *History of depression/anxiety (%)* | 31.3 | 64.3 | 50.0 | NS |
| *Cohabitant causing sleep disruption (%)* | 28.6 | 18.2 | 14.3 | NS |

**Supplementary Table 1. Cohort demographics at 10-year follow up.** N/A=non-applicable. NS=non-significant. Group differences assessed by independent Student's T-test, One-way ANOVA or Chi square/Fisher's exact test. *Relevant medications in controls: beta blocker(n=1), statin(n=2), calcium channel blocker(n=1), ACE inhibitor(n=1). Relevant medications in HD gene carriers: carbamazepine(n=1), SSRI (n=2), statin(n=1), antihistamine(n=1), beta blocker (n=1), amantadine(n=1). †Relevant comorbidities in HD gene carriers: post nasal drip(n=1), fibromyalgia(n=1), perimenopausal symptoms(n=2). Where % and n are incongruent, this reflects isolated cases of missing data polypharmacy, and/or multiple comorbidities.

| ***10-year follow up cohort demographics: Actigraphy participants*** | *Controls* | *Premanifest HD* | *Prodromal/manifest HD* | *P* |
|---|---|---|---|---|
| *N* | 14 | 12 | 7 | N/A |
| *Baseline:10 year follow up interval (years) (mean, SD)* | 9.4(1.3) | 10.5(0.4) | 9.6(1.0) | **0.023** |
| *Age (mean, SD)* | 58.9(18.3) | 53.5(12.5) | 54.1(11.2) | NS |
| *Sex (% male)* | 42.9 | 50.0 | 0.0 | NS |
| *CAG repeat length (mean, SD)* | N/A | 40.3(1.8) | 42.7(2.1) | **0.020** |
| *Predicted years to onset from baseline (mean, SD)* | N/A | 22.7(11.4) | 10.6(2.6) | **0.015** |
| *Years of education (mean, SD)* | 15.3(3.8) | 15.5(2.2) | 13.8(2.3) | NS |
| *In employment (%)* | 57.1 | 75 | 28.6 | NS |
| *Relevant medications (%)** | 21.4 | 33.3 | 28.6 | NS |
| *History of alcohol excess (%)* | 28.6 | 50.0 | 14.3 | NS |
| *History of caffeine excess (%)* | 14.3 | 50.0 | 28.6 | NS |
| *Relevant medical comorbidities (%)†* | 0.0 | 16.7 | 28.6 | NS |
| *History of depression/anxiety (%)* | 28.6 | 66.7 | 28.6 | NS |
| *Cohabitant causing sleep disruption (%)* | 28.6 | 18.2 | 14.3 | NS |

**Supplementary Table 2. Cohort demographics at 10-year follow up, limited to participants who undertook actigraphy.** N/A=non-applicable. NS=non-significant. Group differences assessed by independent Student's T-test, One-way ANOVA or Chi square/Fisher's exact test. *Relevant medications in controls: beta blocker(n=1), statin(n=2), calcium channel blocker(n=1), ACE inhibitor(n=1). Relevant medications in HD gene carriers: carbamazepine(n=1), SSRI (n=2), statin(n=1), antihistamine(n=1), beta blocker (n=1), amantadine(n=1). †Relevant comorbidities in HD gene carriers: post nasal drip(n=1), fibromyalgia(n=1), perimenopausal symptoms(n=2). Where % and n are incongruent, this reflects isolated cases of missing data, polypharmacy and/or multiple comorbidities.

| ***12-year follow up cohort demographics*** | *Controls* | *Premanifest HD* | *Prodromal/manifest HD* | *P* |
|---|---|---|---|---|
| *N* | 10 | 8 | 15 | N/A |
| *Baseline:12 year follow up interval (years) (mean, SD)* | 11.5(1.0) | 12.7(0.3) | 11.6(1.2) | **0.032** |
| *Age (mean, SD)* | 57.3(15.1) | 50.2(10.1) | 59.7(10.7) | NS |
| *Sex (% male)* | 40.0 | 13.3 | 50.0 | NS |
| *CAG repeat length (mean, SD)* | N/A | 40.4(1.6) | 41.5(2.2) | NS |
| *Predicted years to onset from baseline (mean, SD)* | N/A | 27.0(12.1) | 13.4(6.2) | **0.002** |
| *Years of education (mean, SD)* | 15.4(4.1) | 15.9(12.7) | 14.1(1.9) | NS |
| *In employment (%)* | 70.0 | 62.5 | 35.7 | NS |
| *Relevant medications (%)** | 10.0 | 42.9 | 50.0 | NS |
| *History of alcohol excess (%)* | 40.0 | 57.1 | 23.1 | NS |
| *History of caffeine excess (%)* | 20.0 | 57.1 | 38.5 | NS |
| *Relevant medical comorbidities (%)* | 0.0 | 0.0 | 21.4 | NS |
| *History of depression/anxiety (%)* | 30.0 | 62.5 | 64.3 | NS |

**Supplementary Table 3. Cohort demographics at 12-year follow up.** N/A=non-applicable. NS=non-significant. Group differences assessed by independent Student's T-test, One-way ANOVA or Chi square/Fisher's exact test. *Relevant medications in controls: statin(n=1), calcium channel blocker(n=1), ACE inhibitor(n=1). Relevant medications in premanifest HD: SSRI (n=4), SNRI(n=1), antihistamine(n=1), carbamazepine(n=1), beta blocker(n=1), statin(n=2), ACE inhibitor(n=1), rivastigmine(n=1). Where % and n are incongruent, this reflects isolated cases of missing data, polypharmacy and/or multiple comorbidities. Cohabitant status not recorded as no domestic recordings (actigraphy) undertaken at this timepoint.

| *12-year follow up cohort demographics: Polysomnography participants* | *Controls* | *Premanifest HD* | *Prodromal/manifest HD* | *P* |
|---|---|---|---|---|
| *N* | 9 | 5 | 8 | N/A |
| *Baseline:12 year follow up interval (years) (mean, SD)* | 11.4(0.9) | 12.6(0.1) | 12.6(0.1) | **0.015** |
| *Age (mean, SD)* | 56.8(16.0) | 53.3(10.6) | 53.3(10.6) | NS |
| *Sex (% male)* | 44.4 | 60.0 | 12.5 | NS |
| *CAG repeat length (mean, SD)* | N/A | 39.6(1.1) | 39.6(1.1) | NS |
| *Predicted years to onset from baseline (mean, SD)* | N/A | 28.5(15.8) | 16.9(9.4) | NS |
| *Years of education (mean, SD)* | 15.1(4.2) | 14.9(2.2) | 14.8(1.8) | NS |
| *In employment (%)* | 77.8 | 80.0 | 50.0 | NS |
| *Relevant medications (%)** | 11.1 | 40.0 | 37.5 | NS |
| *History of alcohol excess (%)* | 44.4 | 80.0 | 25.0 | NS |
| *History of caffeine excess (%)* | 22.2 | 60.0 | 37.5 | NS |
| *Relevant medical comorbidities (%)* | 0.0 | 0.0 | 12.5 | NS |
| *History of depression/anxiety (%)* | 33.3 | 80.0 | 62.5 | NS |

**Supplementary Table 4. Cohort demographics at 12-year follow up, limited to participants undertaking polysomnography.** N/A=non-applicable. NS=non-significant. Group differences assessed by independent Student's T-test, One-way ANOVA or Chi square/Fisher's exact test. *Relevant medications in controls: statin(n=1), calcium channel blocker(n=1), ACE inhibitor(n=1), beta blocker(n=1). Relevant medications in HD gene carriers: SSRI (n=3), carbamazepine(n=1), statin(n=1), rivastigmine(n=1). Where % and n are incongruent, this reflects isolated cases of missing data, polypharmacy and/or multiple comorbidities. Cohabitant status not recorded as no domestic recordings (actigraphy) undertaken at this timepoint.

| ***Withdrawal analysis among HD gene carriers: 10-year follow up actigraphy*** | *Actigraphy completed* | *Actigraphy omitted* | *P* |
|---|---|---|---|
| *N* | 19 | 9 | N/A |
| *Age at baseline (mean, SD)* | 43.5(11.5) | 45.1(11.7) | NS |
| *Sex (% male)* | 31.6 | 33.3 | NS |
| *CAG repeat length (mean, SD)* | 41.2(2.2) | 41.0(1.5) | NS |
| *Predicted years to onset at baseline (mean, SD)* | 18.2(10.8) | 17.5(8.9) | NS |
| *Years of education (mean, SD)* | 14.8(2.3) | 13.8(2.8) | NS |
| *Relevant medications (%)** | 10.5 | 55.6 | **0.02** |
| *History of alcohol excess (%)* | 36.9 | 25.0 | NS |
| *History of caffeine excess (%)* | 42.1 | 50.0 | NS |
| *Relevant medical comorbidities (%)†* | 21.1 | 0.0 | NS |
| *History of depression/anxiety (%)* | 52.6 | 85.7 | NS |

**Supplementary Table 5. Withdrawal bias analysis among HD gene carriers: actigraphy at 10-year follow up.** N/A=non-applicable. NS=non-significant. Group differences assessed by independent Student's T-test or Chi square/Fisher's exact test. Relevant medications in those completing actigraphy: carbamazepine(n=1), SSRI(n=1). Relevant medications in those omitting actigraphy: SSRI(n=4), SNRI(n=1), benzodiazepine(n=1), beta blocker(n=1), statin(n=1), ACE inhibitor(n=1), calcium channel blocker(n=1). Relevant comorbidities in those completing actigraphy: perimenopausal symptoms(n=2), post nasal drip(n=1), fibromyalgia(n=1). Where % and n are incongruent, this reflects isolated cases of missing data, polypharmacy and/or multiple comorbidities.

| ***Withdrawal analysis among HD gene carriers: 12-year follow up polysomnography*** | *Polysomnography completed* | *Polysomnography omitted* | *P* |
|---|---|---|---|
| *N* | 13 | 15 | N/A |
| *Age at baseline (mean, SD)* | 43.6(10.1) | 44.4(12.7) | NS |
| *Sex (% male)* | 30.8 | 33.3 | NS |
| *CAG repeat length (mean, SD)* | 40.5(2.2) | 41.7(1.7) | NS |
| *Predicted years to onset at baseline (mean, SD)* | 21.3(11.9) | 15.1(7.5) | NS |
| *Years of education (mean, SD)* | 14.8(1.9) | 14.4(3.1) | NS |
| *Relevant medications (%)** | 23.1 | 26.7 | NS |
| *History of alcohol excess (%)* | 46.2 | 20.0 | NS |
| *History of caffeine excess (%)* | 46.2 | 40.0 | NS |
| *Relevant medical comorbidities (%)†* | 7.8 | 27.3 | NS |
| *History of depression/anxiety (%)* | 69.2 | 53.8 | NS |

**Supplementary Table 6. Withdrawal bias analysis among HD gene carriers: polysomnography at 12-year follow up.** N/A=non-applicable. NS=non-significant. Group differences assessed by independent Student's T-test or Chi square/Fisher's exact test. Relevant medications in those who undertook polysomnography(PSG): carbamazepine(n=1), SSRI(n=2), beta blocker(n=1). Relevant medications in those who omitted PSG: SSRI(n=3), SNRI(n=1), benzodiazepine(n=1), statin(n=1), ACE inhibitor(n=1), calcium channel blocker(n=1). Relevant comorbidities in those who undertook PSG: post nasal drip(n=1). Relevant comorbidities in those who omitted PSG: perimenopausal symptoms(n=2), fibromyalgia(n=1) Where % and n are incongruent, this reflects isolated cases of missing data, polypharmacy and/or multiple comorbidities. Cohabitant status not recorded as no domestic recordings (actigraphy) undertaken at this timepoint.

| *Clinical Assessment* | ***Baseline*** | | | ***10-year FU*** | | | | | ***12-year FU*** | | | | |
|---|---|---|---|---|---|---|---|---|---|---|---|---|---|
| | Controls | HD | P | Controls | Premanifest HD | Prodromal/ manifest HD | P | $\eta_p^2$ | Controls | Premanifest HD | Prodromal/ manifest HD | P | $\eta_p^2$ |
| *n* | 21 | 28 | N/A | 14 | 14 | 7 | N/A | . | 9 | 6 | 8 | N/A | . |
| *MOCA* | 27.8(2.2) | 27.4(1.7) | NS | 28.0(2.3) | 27.5(1.7) | 25.9(2.2) | NS | . | 29.0(1.7) | 27.8(1.9) | 27.1(3.4) | NS | . |
| *Trail A* | 30.1(8.9) | 31.3(10.1) | NS | 26.2(6.8)* | 22.3(6.1)*** | 35.3(16.9) | **0.016** | **0.24** | 28.3(9.3)* | 21.3(4.1)** | 40.6(18.0) | **0.007** | **0.39** |
| *Trail B* | 53.7(24.7) | 53.8(20.8) | NS | 62.2(25.5)*** | 53.8(19.9)*** | 133.3(53.0) | **0.002** | **0.52** | 57.8(20.7)* | 41.9(11.4)*** | 94.5(43.4) | **0.001** | **0.54** |
| *HVLT delayed* | 10.2(2.4) | 9.3(2.2) | NS | 10.1(10.7)*** | 10.7(1.6)*** | 5.3(3.0) | **<0.001** | **0.59** | 10.7(2.2) | 9.8(1.3) | 9.1(2.9) | NS | . |
| *SDMT* | 50.5(9.8) | 51.2(9.3) | NS | 49.7(10.7)*** | 52.3(10.3)*** | 30.3(10.3) | **<0.001** | **0.52** | 53.9(12.4)** | 55.3(5.2)** | 38.3(9.9) | **0.006** | **0.40** |
| *MADRS* | 2.6(2.7) | 5.0(7.6) | NS | 2.5(2.3) | 4.7(4.9) | 6.0(4.2) | NS | . | 5.3(4.5) | 6.6(5.2) | 7.3(7.9) | NS | . |
| *UHDRS TMS* | N/A | 0.6(1.3) | N/A | N/A | . | . | . | . | N/A | 1.4(2.7) | 18.3(9.8) | **0.004** | **0.62** |
| *UHDRS TFC* | N/A | 13.0(1.1) | N/A | N/A | . | . | . | . | N/A | 12.7(0.8) | 10.9(2.7) | NS | . |
| *UHDRS DCL* | N/A | 0.2(0.4) | N/A | N/A | 0.4(0.5) | 3.5(0.5) | **<0.001** | **0.94** | N/A | 0.1(0.4) | 3.4(0.8) | **<0.001** | **0.81** |
| *NfL (pg/ml)* | 20.6(9.6) | 30.4(15.1) | NS | . | . | . | . | . | 28.9(12.9)*** | 34.4(16.7)** | 83.0(34.1) | **0.003** | **0.51** |

**Supplementary Table 7. Clinical assessment outcomes.** N/A=non-applicable. NS=non-significant. FU=follow up. Group differences assessed by ANCOVA adjusted for age, sex, CAG repeat length, MADRS depression score, relevant medication use and individual interval between baseline and follow up. Values represent totals or mean(SD) as applicable. *p<0.05, **p<0.01, ***p<0.001 in post hoc Tukey test versus prodromal/manifest HD group. Effect sizes reported where p<0.05 in both pairwise post hoc assessments vs prodromal/manifest HD.

| *Video vs in-person assessment* | ***10-year FU (video)*** | ***12-year FU (in person)*** | ***P*** |
|---|---|---|---|
| *MOCA* | 27.9(1.8) | 28.2(1.8) | NS |
| *Trail A* | 24.3(6.3) | 28.5(10.2) | NS |
| *Trail B* | 65.4(26.8) | 63.7(31.2) | NS |
| *HVLT delayed* | 10.1(2.7) | 10.3(2.1) | NS |
| *SDMT* | 48.9(10.2) | 50.4(11.0) | NS |
| *MADRS* | 4.3(4.4) | 5.2(4.1) | NS |

**Supplementary Table 8. Comparison of clinical assessments undertaken by video call versus in person.** Paired samples Student's T-test applied to n=20 participants who undertook both forms of assessment. Values represent mean(SD). NS=non-significant. FU=follow up.

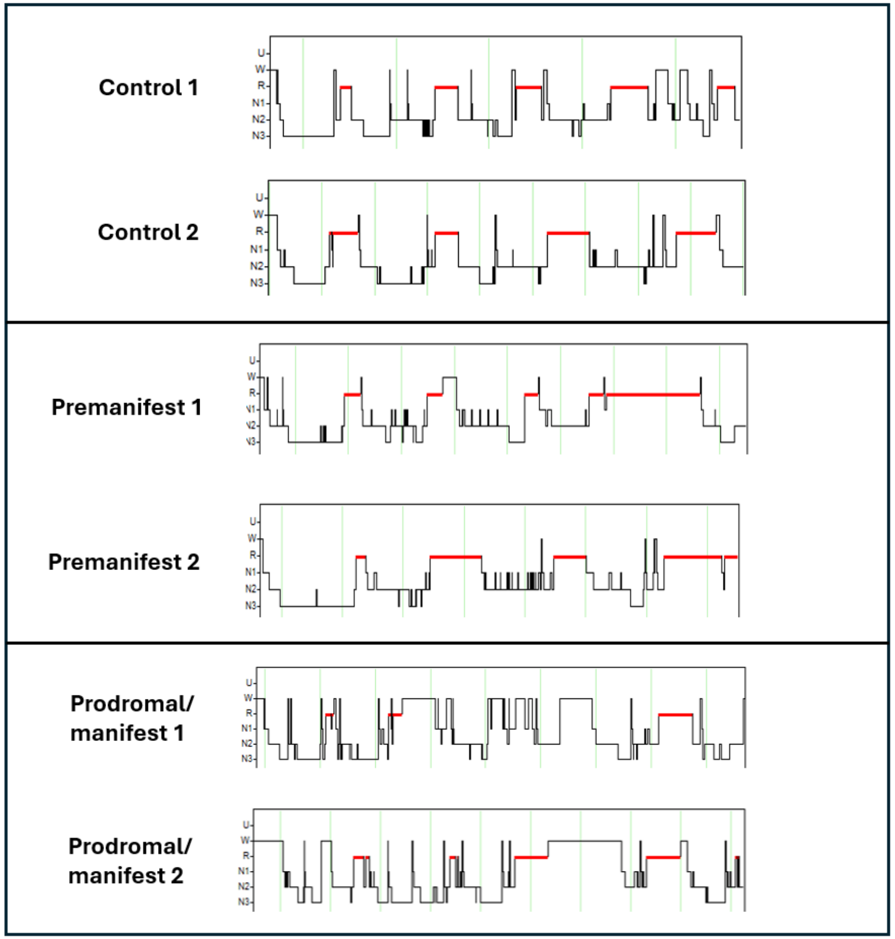


**Supplementary Figure 1. Example PSG hypnograms from six participants at 12-year follow up**, illustrating sleep stage instability and sleep maintenance insomnia in patients with prodromal/manifest HD. U=unscored, W=wake, R=REM (red), N1=stage 1 sleep, N2=stage 2 sleep, N3=slow wave sleep

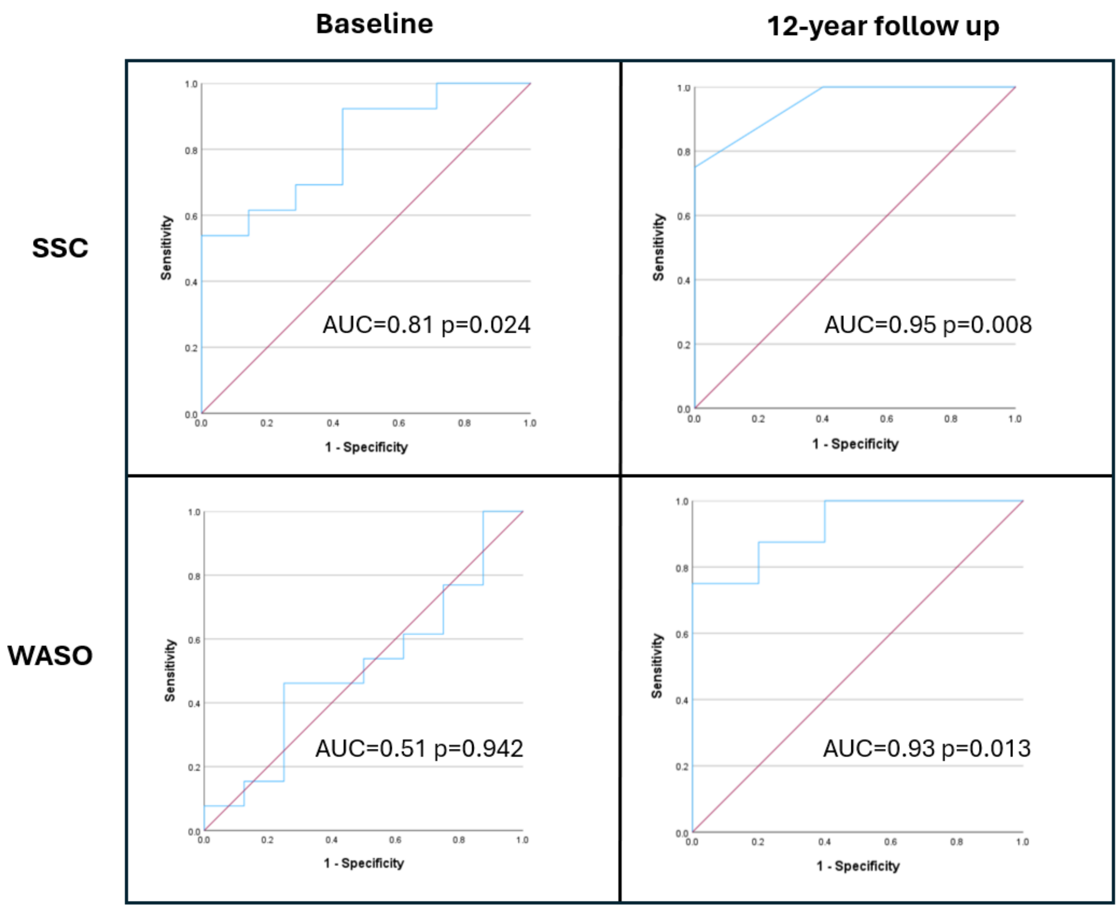


**Supplementary Figure 2. ROC curve analysis** of the ability of SSC and WASO to discriminate phenoconversion outcome among HD gene carriers at study completion, at baseline (left) and follow up (right). AUC= area under curve.
